# Identifying Acute Low Back Pain Episodes in Primary Care Practice from Clinical Notes

**DOI:** 10.1101/19010462

**Authors:** Riccardo Miotto, Bethany L. Percha, Benjamin S. Glicksberg, Hao-Chih Lee, Lisanne Cruz, Joel T. Dudley, Ismail Nabeel

## Abstract

**Background:** Acute and chronic low back pain (LBP) are different conditions with different treatments. However, they are coded in electronic health records with the same ICD-10 code (M54.5) and can be differentiated only by retrospective chart reviews. This prevents efficient definition of data-driven guidelines for billing and therapy recommendations, such as return-to-work options.

**Objective:** To solve this issue, we evaluate the feasibility of automatically distinguishing acute LBP episodes by analyzing free text clinical notes.

**Methods:** We used a dataset of 17,409 clinical notes from different primary care practices; of these, 891 documents were manually annotated as “acute LBP” and 2,973 were generally associated with LBP via the recorded ICD-10 code. We compared different supervised and unsupervised strategies for automated identification: keyword search; topic modeling; logistic regression with bag-of-n-grams and manual features; and deep learning (ConvNet). We trained the supervised models using either manual annotations or ICD-10 codes as positive labels.

**Results:** ConvNet trained using manual annotations obtained the best results with an AUC-ROC of 0.97 and F-score of 0.69. ConvNet’s results were also robust to reduction of the number of manually annotated documents. In the absence of manual annotations, topic models performed better than methods trained using ICD-10 codes, which were unsatisfactory for identifying LBP acuity.

**Conclusions:** This study uses clinical notes to delineate a potential path toward systematic learning of therapeutic strategies, billing guidelines, and management options for acute LBP at the point of care.

## Introduction

Low back pain (LBP) is one of the most common causes of disability in US adults under the age of 45 [1], with 10-20% of American workers reporting persistent back pain [2]. LBP impacts one’s ability to work and affects the quality of life. For example, in 2015 Luckhaupt et al. showed that, from a pool of 19,441 people, 16.9% of workers with any LBP and 19.0% of those with frequent and severe LBP missed at least one full day of work over a period of three months [3]. LBP events also lead to significant financial burden for both individuals and clinical facilities, with combined direct and indirect costs of treatment for musculoskeletal injuries and associated pain estimated to be approximately $213 billion annually [4].

LBP events fall into two major categories: acute and chronic [5]. Acute LBP occurs suddenly, usually associated with trauma or injury with subsequent pain, whereas chronic LBP is often reported by patients in regular checkups and has led to a significant increase in the use of healthcare services over the past two decades. It is very important to differentiate between acute and chronic LBP in the clinical setting as these conditions - as well as their management and billing - are substantively different. Chronic back pain is generally treated with spinal injections [6,7], surgery [8,9], and/or pain medications [10,11], while anti-inflammatories and a rapid return to normal activities of daily living are generally the best recommendations for acute LBP [12].

However, acute and chronic LBP are usually not explicitly separated in electronic health records (EHRs) due to a lack of distinguishing codes. The ICD-10-CM (International Classification of Diseases, Tenth Revision, Clinical Modification) standard only includes the code M54.5 to characterize “Low back pain” diagnosis, and does not provide modifiers to distinguish different LBP acuities [13]. Acuity is usually reported in clinical notes, requiring retrospective chart review of the free text to characterize LBP events, which is time-consuming and not scalable [14]. Moreover, acuity can be expressed in different ways. For example, the text could mention “acute low back pain” or “acute lbp”, but could also simply report “shooting pain down into the lower extremities”, “limited spine range of motion”, “vertebral tenderness”, “diffuse pain in lumbar muscles”, and so on [15]. This variability makes it difficult for clinical facilities and researchers to group LBP episodes by acuity to perform key tasks, such as defining appropriate diagnostic and billing codes; evaluating the effectiveness of prescribed treatments; and deriving therapeutic guidelines and improved diagnostic methods that could reduce time, disability and cost.

This paper is the first to explore the use of automated approaches based on machine learning and information retrieval to analyze free-text clinical notes and identify the acuity of LBP episodes. Specifically, we use a set of manually annotated notes to train and evaluate various machine learning architectures based on logistic regression, n-grams, topic models, word embeddings and convolutional neural networks, and to demonstrate that some of these models are able to identify acute LBP episodes with promising precision. In addition, we demonstrate the ineffectiveness of using ICD-10 codes alone to train the models, reinforcing the idea that they are not sufficient to differentiate the acuity of LBP. Our overall objective is to build an automated framework that can help front line primary care providers in the development of targeted strategies and return-to-work (RTW) options for acute LBP episodes in clinical practice.

## Background and Significance

Primary care providers (PCPs) are commonly the first medical practitioners to assess patient’s musculoskeletal injuries and pain associated with these injuries and are therefore in a unique position to offer reassurance, treatment options, and RTW recommendations catered to the acuity of the injury and pain associated with it. Several studies have documented increases in medication prescriptions and visits to physicians, physical therapists, and chiropractors for LBP episodes [16–18]. Since individuals with chronic LBP seek care and use health care services more frequently than those with acute LBP, increases in health care use and costs for back pain are driven more by chronic than acute cases [19].

A rapid return to normal activities of daily living, including work, is generally the best activity recommendation for acute LBP management [12]. The number of workdays that are lost due to acute LBP can be reduced by implementing clinical practice guidelines in the primary care setting [20]. In previous work, Cruz et al. built a RTW protocol tool for PCPs based on guidelines from the LBP literature [21]. Based on the type of work (e.g., clerical, manual, or heavy) and the severity of the condition, the doctor would recommend RTW options (in partial or full duty capacity) within a certain number of days. The study found that physicians were likely to use this protocol, especially when it was integrated into the EHRs. The protocol was not always used for patients suffering from acute LBP, however, as the research team was unable to quickly identify the acuity using only the structured EHR data (e.g., ICD-10 codes). Acuity information was only available in the progress notes and was thus not incorporated into the automated recommendations. This prevented the research team from providing an accurate feedback to PCPs based on a full picture of the patient’s condition. A similar tool that could incorporate acuity information from notes could provide much more specific recommendations to PCPs that incorporate best practice guidelines for each acuity level. Besides leading to more precise care, this would streamline billing for LBP [22]. Similar needs arise for other musculoskeletal conditions, such as knee, elbow, and shoulder pain, where ICD-10 codes do not differentiate by pain level and acuity [23,24].

Machine learning methods for EHR data processing are enabling improved understanding of patient clinical trajectories, creating opportunities to derive new clinical insights [25,26]. In recent years, the application of deep learning, a hierarchical computational design based on layers of neural networks [27], to structured EHRs has led to promising results on clinical tasks like disease phenotyping and prediction [28–33]. However, a wealth of relevant clinical information remains locked behind clinical narratives in the free text of notes. Natural Language Processing (NLP), a branch of computer science that enables machines to understand and process human language [34] for applications like machine translation [35], text generation [36], and image captioning [37], has been used to parse clinical notes to extract relevant insights that can guide clinical decisions [38]. Recent applications of deep learning to clinical NLP have classified clinical notes according to diagnosis or disease codes [39–41], predicted disease onset [32,42], and extracted primary cancer sites and their laterality in pathology reports [43,44]. However, while deep learning has successfully been applied to analyze clinical notes, traditional methods are still preferable when training data are limited [45,46].

Regardless of the specific methodology, tools based on NLP applied to clinical narratives have not been widely used in clinical settings [31,38], despite the fact that physicians are likely to follow computer-assisted guidelines if recommendations are tied to their own observations [47]. In this paper, we present an NLP-based framework that can help physicians adhere to best practices and RTW recommendations for LBP. To the best of our knowledge, there are no studies to date that have applied machine learning to clinical notes to distinguish the acuity of a musculoskeletal condition in cases where it is not explicitly coded.

## Methods

The conceptual steps of this study are summarized in Figure 1, specifically: dataset composition; text processing; clinical notes modeling; and experimental evaluation. The overall goal was to evaluate the feasibility of automatically identifying clinical notes reporting “acute LBP” episodes.

**Figure 1:**
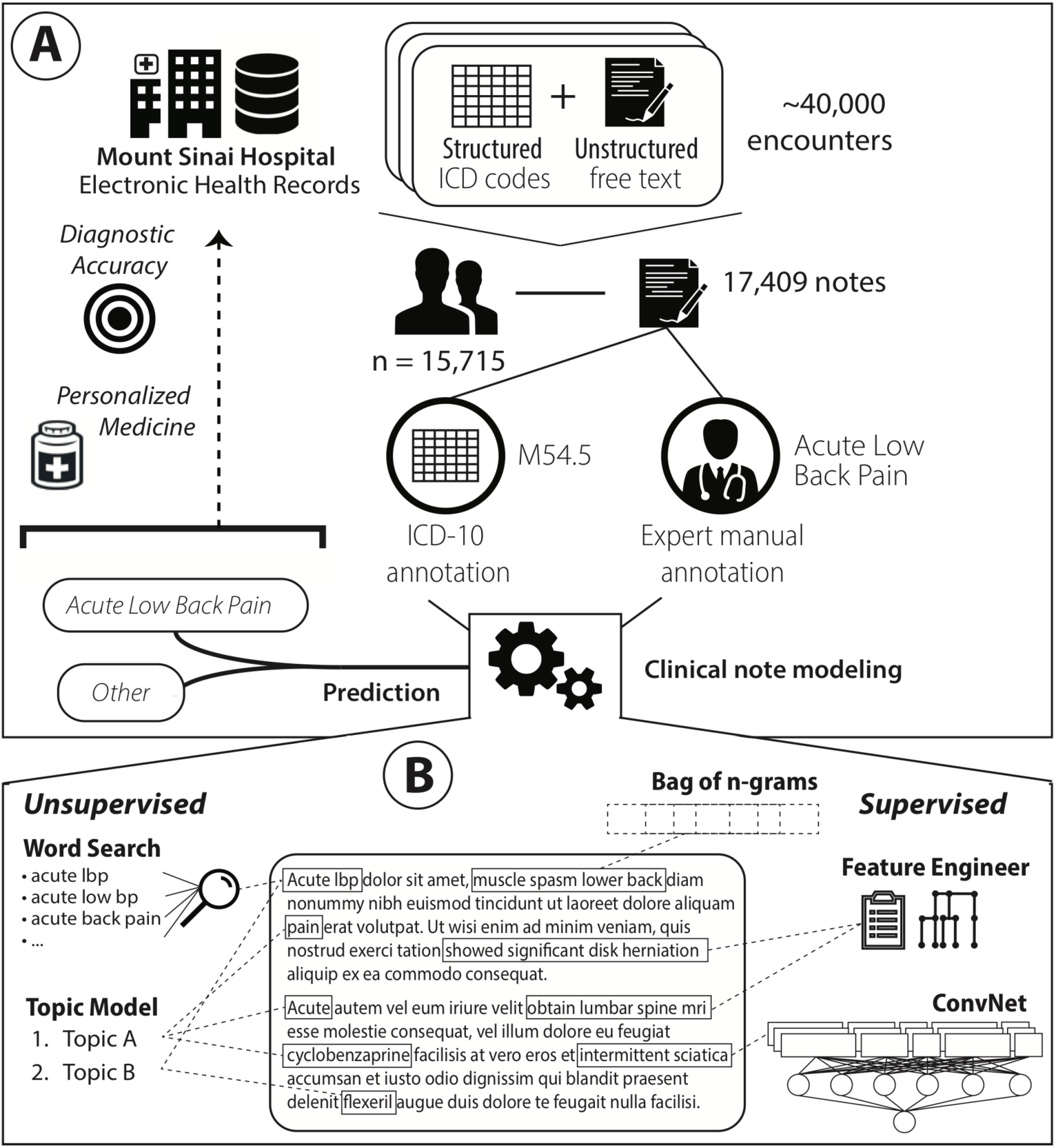
Conceptual framework used to evaluate the use of automated approaches based on machine learning and information retrieval to analyze free-text clinical notes and identify “acute low back pain” episodes.

### Dataset

We used a set of free-text clinical notes extracted from the Mount Sinai data warehouse, made available for use under IRB approval following HIPAA guidelines. The Mount Sinai Health System is an urban tertiary care hospital located in the Upper East Side of Manhattan in New York City. It generates a high volume of structured, semi-structured, and unstructured data as part of its routine healthcare and clinical operations, which include inpatient, outpatient, and emergency room visits. These clinical notes were collected during a previous pilot study evaluating a RTW tool based on EHR data that included nearly 40,000 encounters for 15,715 patients spanning the years 2016 - 2018 and clinical notes written by 81 different providers (Cruz et al. [21]). In that study, we used the published literature to develop a list of guidelines to determine the assessment and management of acute LBP episodes in clinical practice. In particular we used ICD-10 codes as well as other parameters, such as “presenting complaint”, “pre-existing conditions”, “management factors”, “imaging/radiology/test ordered”, and so on, to define and label the acuity of LBP in a clinical encounter. Following these guidelines, 14 individuals (physical medicine and rehabilitation fellows, residents, and medical students) manually reviewed a random set of 4,291 clinical notes associated with these encounters and labeled all “acute low back pain” events. Each note was reviewed by at least two individuals and was further checked by a lead physician researcher if it was marked as ambiguous and/or there was discordance between reviewers. This project leveraged the entire set of clinical notes that were collected in the previous study. In particular, we joined all the progress notes of these encounters under the same initial visit, and we eliminated duplicate, short (less than 3 words), and non-meaningful reports. The final dataset was composed of 17,409 distinct clinical notes, with length ranging from seven to 6,638 words. Of this set, 3,092 notes were manually reviewed in the previous study and 891 of them were annotated as “acute LBP”. The remaining 14,317 notes were not manually evaluated and were related to different clinical domains, including various musculoskeletal disorders and potentially LBP events. In this final dataset, 1,973 notes were also associated to an encounter billed with an ICD10 M54.5 “Low back pain” code.

### Text Processing

Every note in the dataset was tokenized, divided into sentences, and checked to remove punctuation, numbers, and non-relevant concepts such as URLs, emails, dates, etc. Each note was then represented as a list of sentences, with every sentence being a list of lemmatized words represented as one-hot encodings. The vocabulary was composed of all the words appearing at least five times in the training set. The discarded words were corrected to the terms in the vocabulary having the minimum edit distance, i.e., the minimum number of operations required to transform one string into the other [48]. This step reduced the number of misspelled words and prevented the accidental discarding of relevant information; at the same time it also limited the size of the vocabulary to improve scalability [39]. Overall, the vocabulary covering the whole dataset was comprised of 56,142 unique words.

### Clinical Note Modeling

We evaluated different approaches for identifying clinical notes that refer to acute LBP episodes. These included both supervised and unsupervised methods. While we benefited from the use of high-quality manual annotations to train the supervised models, we also investigated alternatives that did not require manual annotation of notes. All of these methods provided straightforward explanations of their predictions, enabling us to validate each model and to identify parts of text and patterns that are relevant to the “acute LBP” predictions.

#### Keyword Search

We searched for a set of relevant keywords in the text. In particular, we looked for “acute low back pain”, “acute lbp”, “acute low bp”, and “acute back pain” and we counted their occurrences in the text. We used NegEx [49] to remove negated occurrences of the keywords. In the evaluation, we refer to this model as “WordSearch”.

#### Topic Modeling

We used topic modeling on the full set of words contained in the notes to capture abstract topics referred to in the dataset [50]. Topic modeling is an unsupervised inference process, in this case implemented using latent Dirichlet allocation [51], that captures patterns of word co-occurrences within documents to define interpretable topics (i.e., multinomial distribution of words) and represent a document as a multinomial over these topics. Every document can then be classified as talking about one or (usually) more topics. Topic modeling is often used in healthcare to generalize clinical notes, improve the automatic processing of patient data, and explore clinical datasets [52–55].

In this study, we assumed that one or more of these topics might refer to acute LBP. In order to discover them, we identified the most likely topics for a set of keywords (i.e., “acute”, “low”, “back”, “pain”, “lbp”, “bp”) and we manually reviewed them to retain only those that seemed more likely to characterize acute LBP episodes (i.e., that included most of the keywords with high probability). We then considered the maximum likelihood among these topics as the probability that a report referred to acute LBP (i.e., “TopicModel” in the experiments).

#### Bag of N-grams

Each clinical note was represented as a bag of n-grams (with n = 1, …, 5), with Term Frequency-Inverse Document Frequency (tf-idf) weights (determined from the corpus of documents). Each n-gram is a contiguous sequence of *n* words from the text. We considered all the words in the vocabulary and filtered the common stop words based on the English dictionary before building all the n-grams. The classification was implemented using Logistic Regression with Lasso (i.e., “BoN-LR”).

#### Feature Engineering

We used the protocol built by Cruz et al. [21] to define acute LBP episodes in the clinical notes. In particular, we used all the concepts described in that guideline, pre-processed them with the same algorithm used for the clinical notes, and built a set of 5,154 distinct n-grams (with n = 1, …, 5), that we refer to as “FeatEng”. We then represented each clinical note as a bag of FeatEng (i.e., we counted the occurrences of only these n-grams in the text), normalized with tf-idf weights, and classified them using Logistic Regression with Lasso (i.e., “FeatEng-LR”).

#### Deep Learning

We implemented an end-to-end deep neural network architecture (i.e., “ConvNet”) that takes as input the full note and outputs its probability of being related to “acute LBP”. The first layer of the architecture maps the words to dense vector representations (i.e, “embeddings”), which attempt to contextualize the semantic meaning of each word by creating a metric space where vectors of semantically similar words are close to each other. We applied word2vec with the skip-gram algorithm to the parsed notes [56] to initialize the embedding of each word in the vocabulary. Word2vec is commonly used with EHRs to learn embeddings of medical concepts from structured data as well as from clinical notes [46,57–59].

The embeddings were then fed to a Convolutional Neural Network (CNN) inspired by the model described by Kim [60] and by Liu *et al*. [42]. This architecture concatenates representations of the text at different levels of abstraction, by essentially choosing the most relevant n-grams at each level. Here, we first applied a set of parallel 1D convolutions on the input sequence with kernel sizes ranging from 1 to 5, thus simulating n-grams with n = 1, …, 5. The outputs of each of these convolutions were then max-pooled over the whole sequence and concatenated to a 5 × *d* dimensional vector, where *d* is the number of 1D convolutional filters. This representation was then fed to sequences of fully connected layers, which learn the interactions between the text features, and finally to a sigmoid layer that outputs the prediction probability.

The n-grams that are most relevant to the prediction, in this architecture, are those that activate the neurons in the max-pooling layer. Therefore, we used the log-odds that the n-gram contributes to the sigmoid decision function [42] as an indication of how much each n-gram influences the decision.

### Evaluation Design

We evaluated all the architectures using a 10-fold cross-validation experiment, with every note appearing in the test set only once. In each training set we used a random 90/10 split to train and validate all the model configurations. As baseline we also report the results obtained by considering as “acute LBP” all the notes associated with the “Low back pain” M54.5 ICD-10 code (i.e., “ICD-10” in the results).

#### Training Annotations

We considered two different sets of annotations as gold standards to train the supervised models. In the first experiment, we used the manually curated annotations provided with the dataset from previous work [21], whereas in the second experiment we trained the models using the ICD-10 codes associated with each note encounter. Both experiments were evaluated using manual annotations. The rationale was to compare the feasibility of identifying acute LBP events when manual annotations are and are not available. We trained the classifier to output *“acute LBP” vs. “other”* because the goal of the project was to identify clinical notes with acute LBP events, rather than discriminate different facets of LBP events (e.g., “chronic LBP” vs. “acute LBP”).

#### Metrics

For all experiments, we report area under the receiver operating characteristic curve (AUC-ROC), micro-precision, recall, F-score, and area under the precision-recall curve (AUC-PRC) [61]. The ROC curve is a plot of true positive rate versus false positive rate found over the set of predictions. F-score is the harmonic mean of classification precision and recall per annotation, where precision is the number of correct positive results divided by the number of all positive results, and recall is the number of correct positive results divided by the number of positive results that should have been returned. The PRC is a plot of precision and recall for different thresholds. The areas under the ROC and PRC curves are computed by integrating the corresponding curves.

#### Model Hyperparameters

The model hyperparameters were empirically tuned using the validation sets to optimize the results with both training annotations.

In the topic modeling method, we inferred topics using the whole training set of documents and 200 topics (derived using perplexity analysis). While seemingly more intuitive, using only the notes associated with the M54.5 “Low back pain” ICD10 code actually produced worse results. For each fold, the most relevant topics associated with acute LBP were manually reviewed and used to annotate the notes.

In the deep learning architecture, we used embeddings with size 300, and full-length notes. We trained word2vec just on the clinical note dataset to initialize embeddings. Pre-initializing the embeddings with a general-purpose corpus did not lead to any improvement. Each CNN had 200 filters and used a ReLu activation function. We added two fully connected layers of size 600 following the CNNs with ReLu activations and batch normalization. Dropout values across the layers were all set to 0.5. The architecture was trained using cross-entropy loss with the Adam optimizer for five epochs and batch size 32 (learning rate = 0.001). The classification thresholds for precision, recall, and F-score were found by ranging the value from 0.1 to 1, with 0.1 increments, and retaining, for each model, the value leading to the best results on the validation set.

## Results

Table 1 and Figure 2 show the average results of the 10-fold cross-validation experiment for all the models considered. The best results were obtained by ConvNet when trained with the manual annotations. While this is not entirely surprising given the success of deep learning for NLP when high-quality annotations and a large amount of data (i.e., on the order of millions of training examples) are available, this was not certain in this domain where the training dataset was much smaller. As expected, the results obtained by the baseline and by training the models using the ICD-10 codes were not as good, confirming that the M54.5 ICD-10 code is not a sufficient indicator of acute LBP. TopicModel leads to similar performance but provides a more intuitive and potentially effective way for exploring the collection, extracting meaningful patterns that are related to acute LBP episodes (see Figure 3). While this approach might not be robust enough for clinical application, a refined and manually curated version of TopicModel promises to allow an efficient pre-filtering of clinical reports that can speed up the manual work required to annotate them. On the contrary but as expected, WordSearch performed poorly as the condition is mentioned in too many different ways across the text and simple keywords were not sufficient.

**Table 1:** Classification results in identifying clinical notes with “acute LBP” episodes in terms of Precision, Recall, F-score, Area Under the ROC (AUC-ROC) and Precision-Recall (AUC-PRC) curves. Results are averaged over the 10-fold cross validation experiment. We compared different supervised and unsupervised strategies: keyword search (“WordSearch”); topic modeling (“TopicModel”); logistic regression with bag-of-n-grams (“BoN-LR”) and manual features (“FeatEng-LR”); and deep learning (“ConvNet”). The supervised models (i.e., BoN-LR, FeatEng-LR and ConvNet) were trained using manual annotations or M54.5 ICD-10 codes. The “ICD-10” baseline simply considered as “acute LBP” all the notes associated with the generic M54.5 “Low back pain” ICD-10 code.

|  |  | Precision | Recall | F-score | AUC-ROC | AUC-PRC |
| --- | --- | --- | --- | --- | --- | --- |
| *Baseline* | ICD-10 | 0.32 | 0.68 | 0.41 | 0.81 | 0.42 |
| *Unsupervised Methods* | WordSearch | **0.71** | 0.03 | 0.06 | 0.52 | 0.40 |
|  | TopicModel | 0.44 | 0.58 | 0.50 | 0.92 | 0.46 |
| *Trained with the M54.5 ICD-10 Code* | BoN-LR | 0.50 | 0.70 | 0.59 | 0.83 | 0.42 |
|  | FeatEng-LR | 0.47 | 0.59 | 0.52 | 0.88 | 0.41 |
|  | ConvNet | 0.55 | 0.68 | 0.61 | 0.89 | 0.46 |
| *Trained with Manual Annotations* | BoN-LR | 0.53 | 0.64 | 0.58 | 0.93 | 0.56 |
|  | FeatEng-LR | 0.58 | 0.66 | 0.62 | 0.93 | 0.58 |
|  | ConvNet | 0.65 | **0.73** | **0.70** | **0.98** | **0.72** |

**Figure 2:**
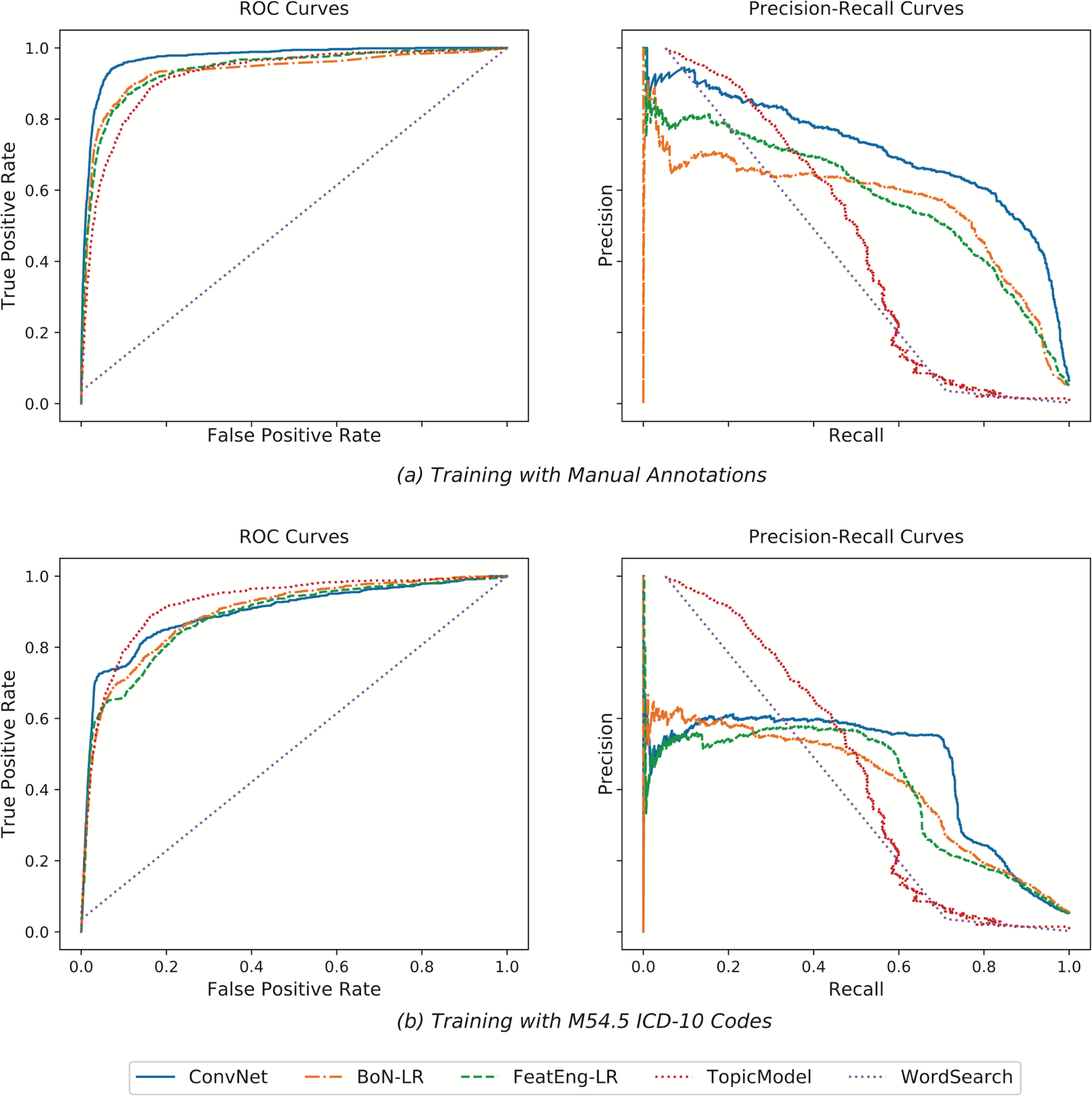
ROC and Precision-Recall curves obtained when using as training data for BoN-LR, FeatEng-LR and ConvNet the manual annotations (a) and the M54.5 ICD-10 codes (b). ConvNet trained using the manual annotations obtained the best results. In the absence of manual annotations to use for training, TopicModel worked better than methods trained using ICD-10 codes, which proved not to be a good indicator to identify acuity in LBP episodes.

**Figure 3:**
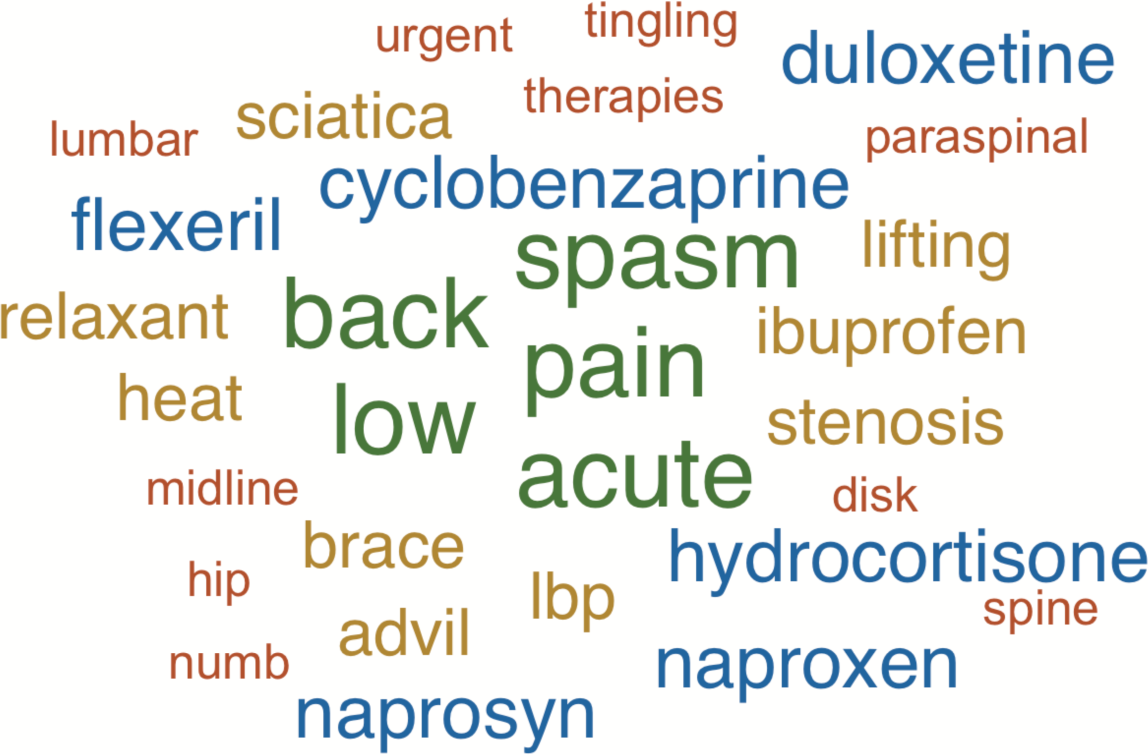
Representative “acute LBP”-related topic derived by averaging the word likelihood in all the relevant topics (i.e., that were manually verified) inferred across the 10-fold cross-validation experiment. We report the top 30 words, with the biggest words being the most relevant. As it can be seen, most of the words are indeed related to acute LBP, including several medications that are usually prescribed to treat inflammation and pain (e.g., Cyclobenzaprine, Flexeril, Advil). A manually refined version of TopicModel can help pre-filtering the notes in an intuitive semi-automatic way, promising to speed up the manual annotation process.

Figure 4 reports the classification results in terms of AUC-ROC and AUC-PRC when randomly subsampling the “acute LBP” manual annotations in the training set. We found that ConvNet always outperforms the other methods based on LR as well as TopicModel. In addition, we notice that using just 30% of the manual annotations (i.e., 70 clinical notes) already leads to better results than using ICD-10 codes as training data. This is a particularly interesting insight as it shows that only minimal manual work is required in order to achieve good classifications; these can then be further improved by adding automatically annotated notes to the model (after manual verification) and retraining.

**Figure 4:**
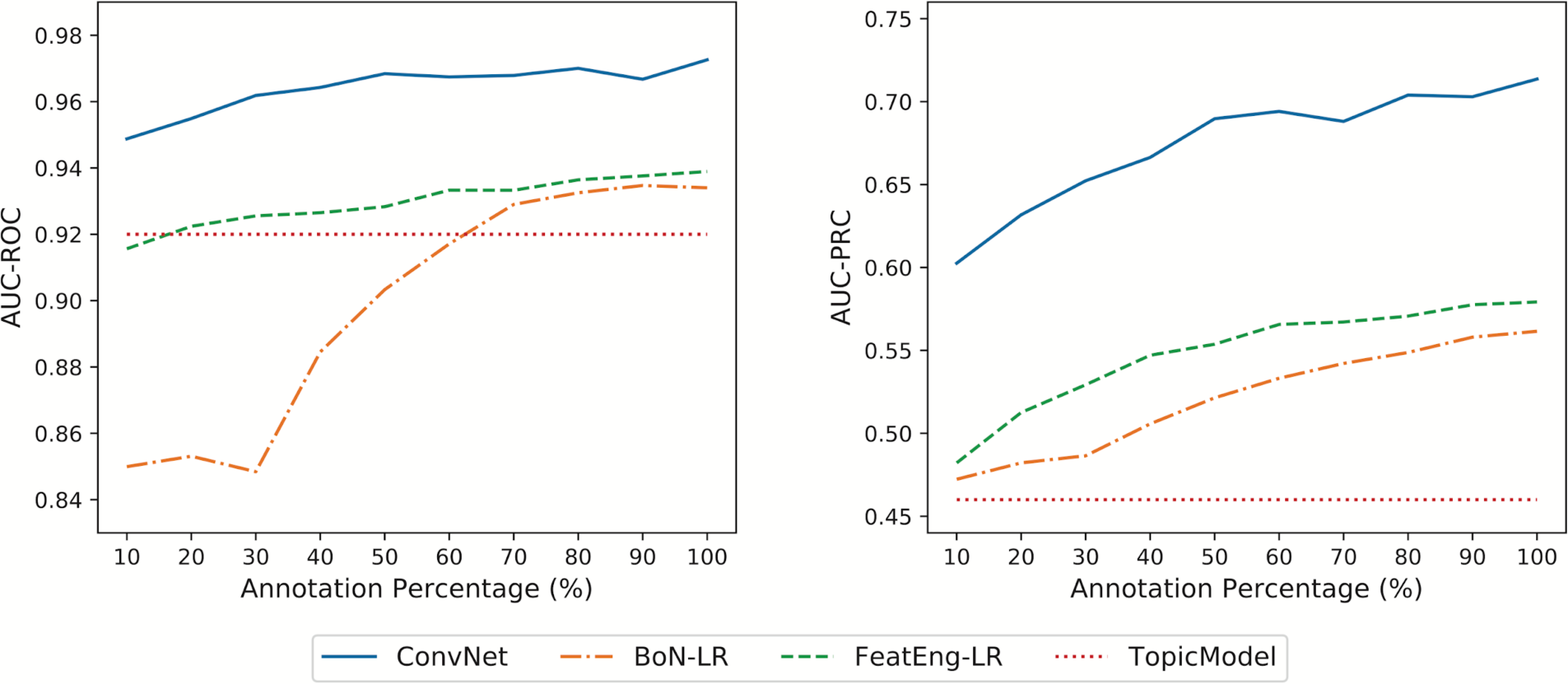
Area under the ROC (AUC-ROC) and Precision-Recall (AUC-PRC) curves obtained when training the supervised models using random sub-samples of the manual annotations. TopicModel is reported as reference baseline. ConvNet obtained satisfactory results when trained using less manually annotated documents, showing robustness and scalability to the gold standard.

Figure 5 highlights the distributions of the classification scores (predicted probability of the label “acute LBP”) derived by several supervised models (trained with manual annotations) and TopicModel. ConvNet shows clear separation between acute LBP notes and the rest of the dataset. In particular, all acute LBP notes had scores greater than 0.2, with 82% of them (i.e., 727 notes) having scores greater than 0.5. On the contrary, only 347 controls had scores greater than 0.5, meaning that only a few notes were highly likely to be misclassified. Similarly, TopicModel had no controls with scores greater than 0.7 and all “acute LBP” notes had scores greater than 0.2.

**Figure 5:**
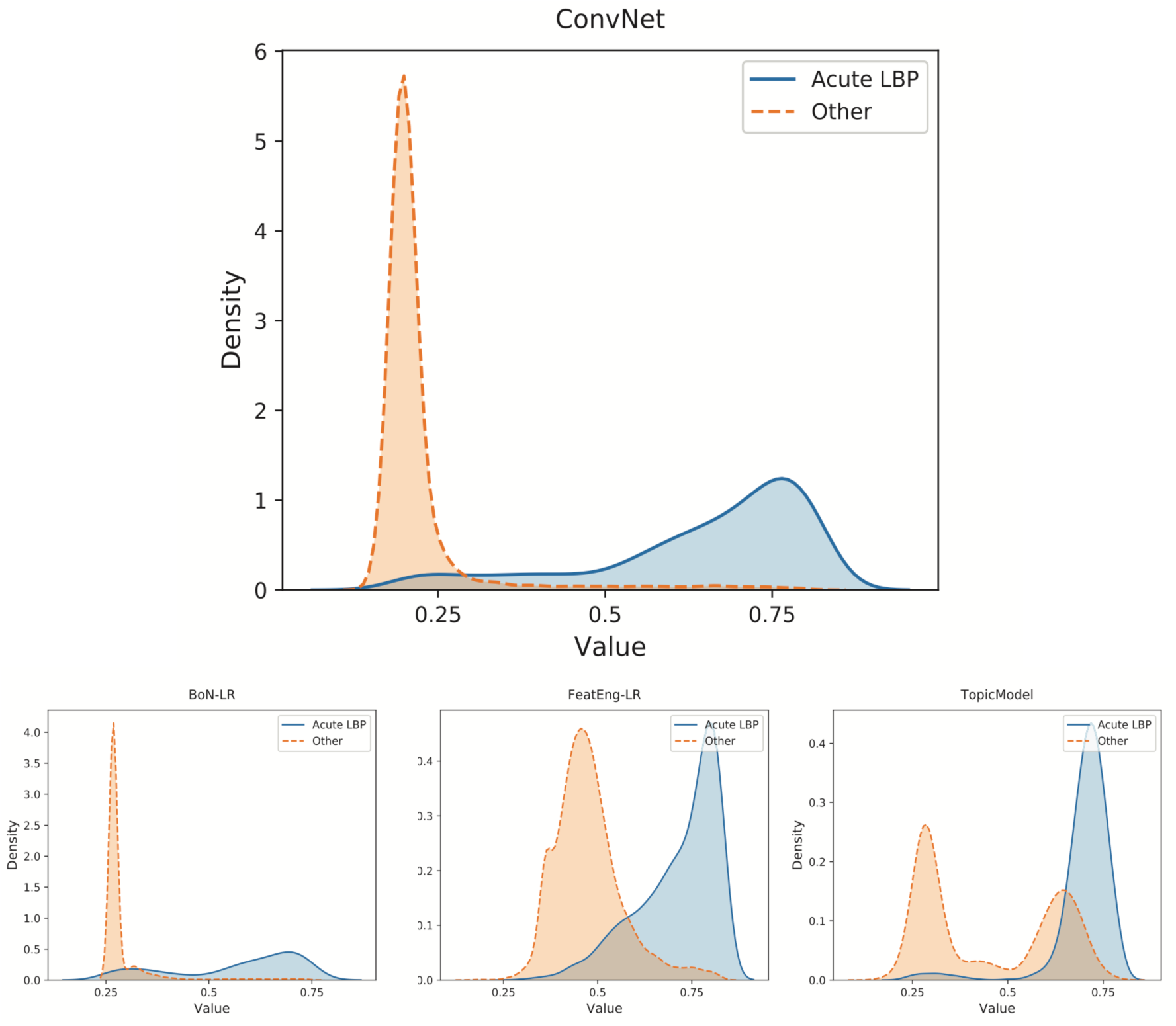
Representation of the probability distribution of the scores obtained by BoN-LR, FeatEng-LR, ConvNet (trained with manual annotations) and TopicModel. ConvNet led to good separation between “acute LBP” clinical notes and all the other documents. In the other cases, such separation is not as clear, explaining the worse classification results obtained by those models.

Finally, Table 2 summarizes some of the n-grams driving the “acute LBP” predictions obtained by ConvNet (trained with manual annotations) across the experiments. While some of these are obvious and refer to the disease itself (e.g., “acute lbp”), others refer to medications (e.g., “prescribed muscle relaxant”, “flexeril”), and recommendations (e.g., “rtw full duty quick”). Given their clinical meaning and relevance, all these patterns can be further analyzed and reviewed to potentially drive the development of guidelines for, e.g., treatment and RTW options.

**Table 2:**
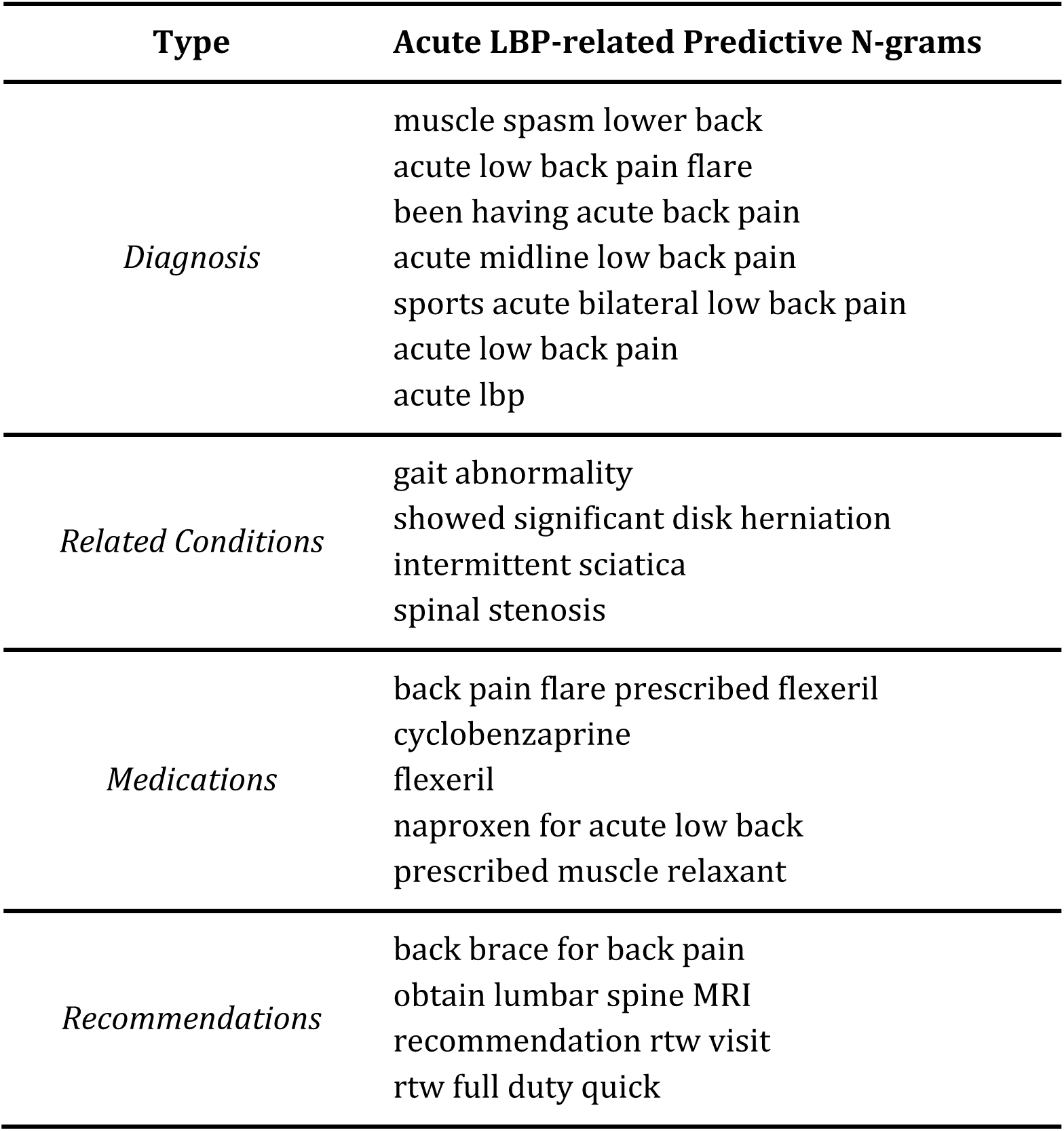
Examples of n-grams that were relevant in identifying “acute LBP” notes when using ConvNet trained with manual annotations. The n-grams relevance was determined by analyzing the neurons of the CNNs activating the max-pooling layers and their log-odds to contribute to the final output. Log-odds were filtered per notes and then averaged over all the notes and evaluation folds.

| Type | Acute LBP-related Predictive N-grams |
| --- | --- |
| *Diagnosis* | muscle spasm lower back<br>acute low back pain flare<br>been having acute back pain<br>acute midline low back pain<br>sports acute bilateral low back pain<br>acute low back pain<br>acute lbp |
| *Related Conditions* | gait abnormality<br>showed significant disk herniation<br>intermittent sciatica<br>spinal stenosis |
| *Medications* | back pain flare prescribed flexeril<br>cyclobenzaprine<br>flexeril<br>naproxen for acute low back<br>prescribed muscle relaxant |
| *Recommendations* | back brace for back pain<br>obtain lumbar spine MRI<br>recommendation rtw visit<br>rtw full duty quick |

## Discussion

In this work we evaluated the use of several machine learning approaches to identify acute LBP episodes in free text clinical notes in order to better personalize the treatment and management of this condition in primary care. The experimental results showed that it is possible to extract acute LBP episodes with promising precision, especially when at least some manually curated annotations are available. In this scenario, ConvNet, a deep learning architecture based on CNNs, significantly outperformed other shallow techniques based on bag-of-n-grams and logistic regression, opening the possibility to boost performances using more complex architectures from current research in the NLP community. The implemented deep architecture also provides an easy mechanism to explain the predictions, leading to informed decision support based on model transparency [62,63] and the identification of meaningful patterns that can drive clinical decision making. If no annotations are available, experiments showed that the use of topic modeling is preferred to training a classifier using only the M54.5 ICD-10 codes (i.e., “Low back pain”) associated with the clinical note encounter, which proved to be a poor indicator to discriminate LBP episodes. In addition, the topics identified can serve as an intuitive tool to inform guidelines and recommendations as well as to pre-filter the documents and reduce the manual work required to annotate the notes. The proposed framework is inherently domain agnostic and does not require any manual supervision to identify relevant features from the free-text. Therefore, it can be leveraged in other musculoskeletal condition domains where acuity is not expressed in the ICD-10/diagnostic codes, such as knee, elbow, and shoulder pain.

### Potential Applications

Medical care decisions are often based on heuristics and manually derived rule-based models constructed on prior knowledge and expertise [64]. Cognitive biases and personality traits, such as aversion to risk or ambiguity, overconfidence, and the anchoring effect, may lead to diagnostic inaccuracies and medical errors resulting in mismanagement or inadequate utilization of resources [65]. In the LBP domain, this may lead to: delays in finding the right therapy and assisting in the return of patients to normal activities; increase in the risk of transitioning the condition from acute to chronic; creating discomfort for patients; and increasing the economic burdens on clinical facilities to adequately treat and manage this patient population. Deriving data-driven guidelines for treatment recommendations can help in reducing these cognitive biases and personality traits, leading to more consistent and accurate decisions. In this scenario, the proposed frameworks integrate seamlessly with the RTW tool proposed by Cruz et al. [21] by including acuity-relevant information in the clinical notes and addressing one of the limitations of that study (i.e., recommending the RTW tool at the point of care by accurately identifying condition as acute LBP). Similarly, an understanding of the patterns driving the predictions can lead to the development of new and improved treatment strategies for various types of injuries, which can be presented to the clinicians at the time of patient encounter to help them with better management of the condition. While physicians will continue to have autonomy in determining optimal care pathways for their patients, the recommendations provided by the supporting framework will be useful to systematize and support their activities within the realm of the busy clinical practice. Posterior analysis of the clinical notes to infer acute LBP episodes can also help in assigning the proper diagnostic and billing codes for the encounter. In a foreseeable future scenario where clinical observations are automatically transcribed via voice and EHRs are processed in real-time, an automated tool that identifies acuity information could also improve the accuracy of diagnosis and billing in real-time, with no need to wait for posterior evaluations.

### Limitations

This work evaluated the feasibility of using machine learning to identify acute LBP episodes in clinical notes. Therefore, we compared different types of models (shallow vs. deep) and learning frameworks (unsupervised vs. supervised) to identify the best directions for implementation and deployment in real clinical settings. While several of the architectures evaluated in this work obtained promising results, more sophisticated models are likely to improve these performances, especially in the deep learning domain. For example, algorithms based on attention models [66], BERT [67], or XLNet [68] have shown encouraging results on similar NLP tasks and are likely to obtain better results in this domain as well. In this work we only focused on processing clinical notes; however, embedding structured EHR data, especially medications, imaging studies and/or lab tests, into the method should improve the results.

The dataset of clinical notes used in this study originated from a geographically diverse set of primary care clinics serving the New York population across the NY metro area over a limited period (i.e., 2016-2018). Providers were enrolled and randomized into the study on a rolling basis, with the number of encounters for LBP varying for each individual provider, based on his/her own practice. The majority of the primary care providers were assistant professors serving on the front lines. No specialists were included in the initial study as the pilot project was only geared towards the primary care providers. Consequently, the results of this study might not be applicable to specialty care practice.

### Future Work

The classification of LBP episodes as acute or chronic at the point of care level within primary care practice is imperative for a RTW tool to be effectively used to render evidence-based guidelines. At this time, we plan to classify a large set of notes, derive patterns related to acute LBP and extend the tool proposed by Cruz et al. [21] according to them. We also plan to identify cases where the RTW tool can be easily deployed based on EHR integration in the clinical domain. Second, we will begin to address some of the methodological limitations of this study to optimize performance and evaluate its generalizability outside primary care. Finally, we aim to evaluate the feasibility of this type of approach for other musculoskeletal conditions, in particular, shoulder and knee pain.

## Conclusions

This article demonstrates the feasibility of using machine learning to automatically identify acute LBP episodes from clinical reports using only unstructured free text data. In particular, manually annotating a set of notes to use as a gold standard can lead to effective results, especially when using deep learning. Topic modeling can help in speeding up the annotation process, initiating an iterative process where initial predictions are validated and then used to refine and optimize the model. This approach provides a generalizable framework for learning to differentiate disease acuity in primary care, which can more accurately and specifically guide the diagnosis and treatment of LBP. It also provides a clear path toward improving the accuracy of coding and billing of clinical encounters for LBP.

## Data Availability

The data used in this study is not publicly available.

## Acknowledgements

I.N. and L.C. would like to thank the Pilot Projects Research Training Program of the NY and NJ Education and Research Center (ERC), National Institute for Occupational Safety and Health, for their funding (grant # T42 OH 008422). R.M. would like to thank the support from the Hasso Plattner Foundation and a courtesy GPU donation from NVIDIA.

## Contributions

R.M. and I.N. initiated the idea and wrote the article; I.N. collected the data and provided clinical support; R.M. conducted the research and the experimental evaluation; B.L.P. advised on evaluation strategies and refined the article; B.S.G., H.L., and L.C. refined the article; J.T.D. supported the research. All the authors edited and reviewed the manuscript.

## Abbreviations

AUC-PRC: Area Under the Precision-Recall Curve
AUC-ROC: Area Under the Receiver Operating Characteristic Curve
BoN: Bag of N-grams
CNN: Convolutional Neural Network
EHR: Electronic Health Record
HIPAA: Health Insurance Portability and Accountability Act
ICD-CM: International Statistical of Diseases, Clinical Modification
IRB: Institutional Review Board
LBP: Low Back Pain
LR: Logistic Regression
NLP: Natural Language Processing
NY: New York
PCP: Primary Care Provider
RTW: Return To Work
TF-IDF: Term Frequency-Inverse Document Frequency

